# Structural Brain Age Trajectories in Antipsychotic-naive First Episode Psychosis

**DOI:** 10.64898/2026.05.25.26353865

**Authors:** Alejandro Roig-Herrero, Shona M. Francey, Brian O’Donoghue, Barnaby Nelson, Laura K.M. Han, Hok Pan Yuen, Andrew Thompson, Kelly Allott, Mario Alvarez-Jimenez, Susy Harrigan, Christos Pantelis, Stephen J Wood, Vanessa Cropley, Patrick McGorry, Alex Fornito, Vicente Molina, Rodrigo de Luis García, Sidhant Chopra

**Affiliations:** Orygen, Parkville, Australia; Centre for Youth Mental Health, The University of Melbourne, Melbourne, Australia; Turner Institute for Brain and Mental Health, School of Psychological Sciences, Monash University, Clayton, Australia; Monash Biomedical Imaging, Monash University, Clayton, Australia; Wu Tsai Institute, Department of Neuroscience, Yale University, New Haven, USA; Department of Psychology, Yale University, New Haven, USA; St Vincent’s University Hospital, Elm Park, Dublin 4, Ireland; School of Medicine, University College Dublin, Ireland; Department of Psychiatry, Brain Health Institute, Rutgers University, Piscataway, NJ, USA; Department of Psychiatry, University of Melbourne, Carlton South, Victoria, Australia; Monash Institute of Pharmaceutical Sciences (MIPS), Monash University, Parkville, Victoria, Australia; Florey Institute of Neuroscience and Mental Health, The University of Melbourne, Parkville, Victoria, Australia; Western Centre for Health Research & Education, University of Melbourne & Western Health, Sunshine Hospital, Furlong Road, St Albans, Victoria, Australia; School of Psychology, University of Birmingham, UK; Faculty of Medicine, University of Valladolid; Psychiatry Service, Clinical Hospital of Valladolid, Valladolid, Spain; Biomedical Research Institute of Valladolid (IBioVall), Spain; Imaging Processing Laboratory, University of Valladolid, Valladolid, Spain; Centre for Mental Health and Brain Sciences, School of Health Sciences, Swinburn University of Technology, Victoria, Australia; Centre for Mental Health, School of Population and Global Health, The University of Melbourne; Department of Psychiatry, Amsterdam UMC location Vrije Universiteit Amsterdam, Amsterdam, The Netherlands; Amsterdam Neuroscience, Mood, Anxiety, Psychosis, Sleep & Stress program, Amsterdam, The Netherlands; Amsterdam Public Health, Mental Health program, Amsterdam, The Netherlands

**Keywords:** Brain Age, First Episode of Psychosis, Longitudinal, MRI, Structural MRI, Schizophrenia

## Abstract

**Background:** Psychotic disorders such as schizophrenia have been associated with older-appearing brain structure, commonly quantified using the brain-age paradigm. However, it remains unclear whether these alterations are present at illness onset and whether antipsychotic treatment modifies their trajectory.

**Methods:** In this study, 61 (28 females and 33 males) antipsychotic-naïve people with first-episode psychosis were randomised to receive either a second-generation antipsychotic (risperidone or paliperidone) or placebo over a 6-month treatment period, alongside intensive psychosocial therapy. A healthy control group (n = 27, 17 females, 10 males) was also recruited. Structural MRI scans were collected at baseline, 3 months, and 12 months. Brain age was estimated using two pretrained and validated models (Pyment and CentileBrain).

**Results:** Brain-predicted age difference (brain-PAD) did not differ between patients and healthy controls at baseline (F_(1,80)_ = 1.30; p = 0.26). There were also no significant effects of time, treatment group (antipsychotic, placebo, healthy control), or their interaction on brain-PAD across the first year (all p > 0.26). Findings were consistent across both brain-age models, and brain-PAD was not associated with clinical and lifestyle measures.

**Conclusion:** These findings suggest that altered structural brain ageing is not evident during the earliest stages of psychosis and is not modified by early antipsychotic exposure over the first year of illness. Longer follow-up and approaches that account for illness heterogeneity may be needed to clarify when brain-age alterations emerge in psychotic disorders.

## Introduction

Psychotic disorders, including schizophrenia, have increasingly been conceptualised as conditions marked by accelerated biological ageing. Several biomarkers of system and organ function have shown this pattern, including telomere erosion, retinal degeneration, and synaptic function (1–4). In the brain, structural Magnetic Resonance Imaging (MRI) provides a noninvasive way to quantify age-related variation and has enabled the reliable estimation of “brain age” as an individual-specific marker of apparent neurobiological aging (5). What remains unclear is whether older-appearing brain structure is detectable during the very early stages of psychotic illness, before antipsychotic exposure, and whether antipsychotic treatment accelerates or attenuates brain age trajectories.

Brain age is typically estimated using a machine learning model trained to predict chronological age from neuroimaging data in healthy individuals. The difference between an individual’s actual age and the predicted age, commonly termed ‘brain-predicted age difference’ (brain-PAD), serves as an indicator of structural brain health. Meta-analyses in schizophrenia consistently show elevated brain-PAD relative to controls, indicative of accelerated ageing (6–8), and this effect appears robust across differences in algorithms and input features (9–21). However, in chronic schizophrenia, elevated brain-PAD may partly reflect cumulative illness burden and non-specific modifiers of brain aging, such as obesity/metabolic morbidity, adverse lifestyle factors and long-term antipsychotic exposure, rather than specific pathophysiology alone (10,18,22–24). Examining individuals during first-episode psychosis (FEP) can allow us to disentangle whether older-appearing brain structure is already present near onset, before chronicity and treatment effects exert their influence.

Findings in FEP samples exposed to antipsychotic medications have been mixed, with some cross-sectional studies reporting higher brain-PAD than controls (11,12,22,25–30), albeit with smaller effects compared to those seen in schizophrenia (6), and often on the order of less than a year of brain-PAD difference in comparison to healthy controls. Other cross-sectional studies have not found differences (31,32), and the two studies that have examined antipsychotic naïve patients have generally reported higher brain-PAD compared to controls (29,30).

Longitudinal naturalistic studies that examine people with schizophrenia have reported a progressive increase in brain-PAD (15,18), with the steepest changes occurring during the first five years of treatment engagement (18). However, this latter finding has not been replicated in follow-up studies of FEP samples (29,30,33), with only one study even reporting a reduction in fractional anisotropy–based brain-PAD within the first five months following the initiation of antipsychotic treatment (29). These inconsistent findings in FEP may be related to variable antipsychotic exposure, with several (13,18,26,33), but not all (10,12,23,24,32), studies demonstrating that higher exposure is associated with a larger brain-PAD.

The effect of antipsychotic medication remains particularly difficult to disentangle because of the ubiquitous and near-immediate treatment of patients and the effect of confounding by indication, where higher doses of antipsychotic medication (30,33) are often given to individuals with more severe symptoms. Therefore, it remains unclear whether antipsychotic treatment alters brain aging trajectories. The optimal way to disentangle these effects *in vivo* is to conduct a clinical trial where antipsychotic-naïve patients experiencing FEP are randomised to receive active antipsychotic medication or a placebo. This design can test several distinct hypotheses about the differential contributions of illness and antipsychotics to brain aging in the earliest stages of illness (Figure 1). We recently completed such a triple-blind placebo-controlled trial with longitudinal structural MRI, including a healthy control group (35,36). In this same cohort, we have previously reported antipsychotic medication-related expansion of the basal ganglia (36), as well as illness-related thinning of the cortex (37). The primary aim of the current work is to determine whether accelerated brain ageing is present during the very early stages of psychotic illness, prior to medication initiation, and to examine the differential effects of antipsychotic medication and illness progression during the first year of illness.

**Figure 1.**
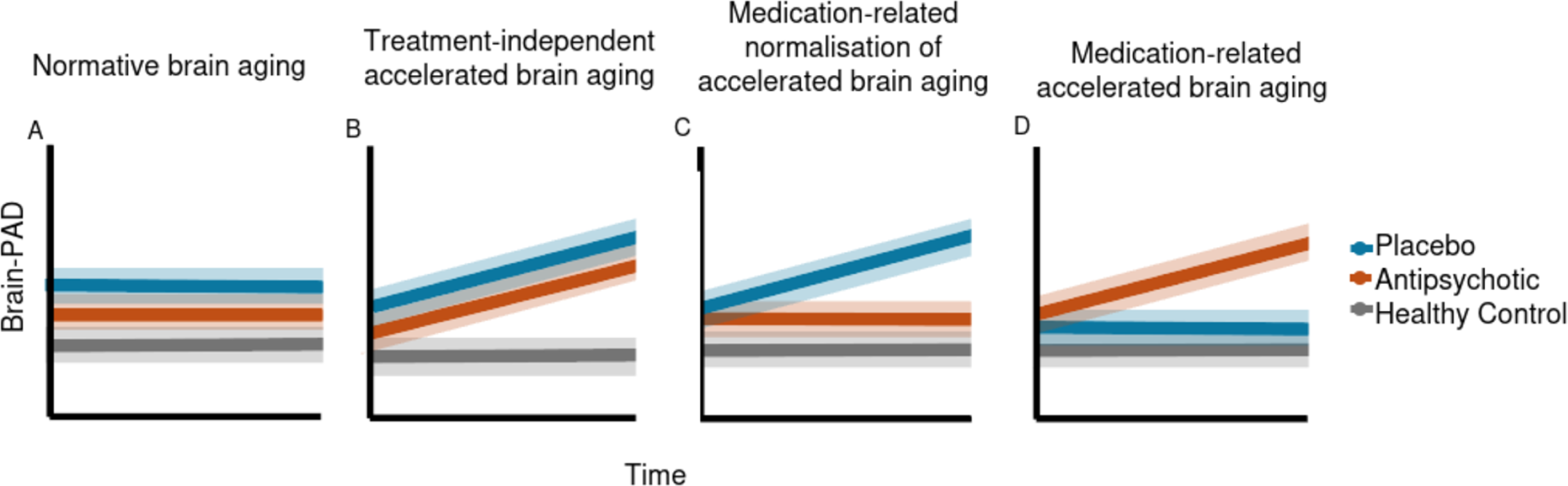
Schematic of expected results under different hypotheses. A) No difference in brain-predicted age difference (Brain-PAD) trajectory between the groups over time would support normative brain aging in FEP, unaffected by medication or illness. B) If both treatment groups show a consistent deviation compared to the control group, this would reflect abnormal brain ageing, unaffected by treatment. C) A deviation in the placebo group trajectory, compared to the medication and control group, would represent a medication-related rescue or normalisation of brain ageing. D) A deviation in medication group trajectory, compared to the placebo and control group, would represent a mediation-related alteration of brain ageing. Note that these hypotheses are for the longitudinal analysis; brain-PAD is set similarly at baseline for simplicity, but may differ between patients and healthy controls. Likewise, deviations from the control group are shown as increased in brain-PAD, where it is possible that decreases may also be detected.

## Methods and Materials

### Study design

We used data from a triple-blind, placebo-controlled longitudinal MRI study which formed part of a larger clinical trial (38). Patients were randomized to one of two groups: one given antipsychotic medication (risperidone [n=25, starting dose = 1mg] or paliperidone [n=5, starting dose = 3mg]) and the other given a placebo, with both groups given intensive psychosocial therapy (38). To reflect real-world clinical practice, the starting dose was then increased by the blinded treating clinician according to clinical response. The same procedure was followed for participants receiving a placebo, who received a pill that was identical in taste, appearance, and packaging to the active medication. A third healthy control group that received no intervention was also recruited. MRI and clinical assessments were conducted at baseline, 3 months, and a final follow-up at 12 months. The randomization phase of the study terminated at 6 months, and treatment as usual continued, so in practice either treatment group could have received antipsychotic medication and ongoing psychosocial interventions between 6 and 12 months into the study. To clearly disentangle the effects of illness from antipsychotics in our analyses, we only included patients in the placebo group who had no exposure to any antipsychotic medication throughout their participation in the first year of the study. This sample has been used in previous research, and further detailed information can be found in Table 1, Supplement and elsewhere (36).

**Table 1.**
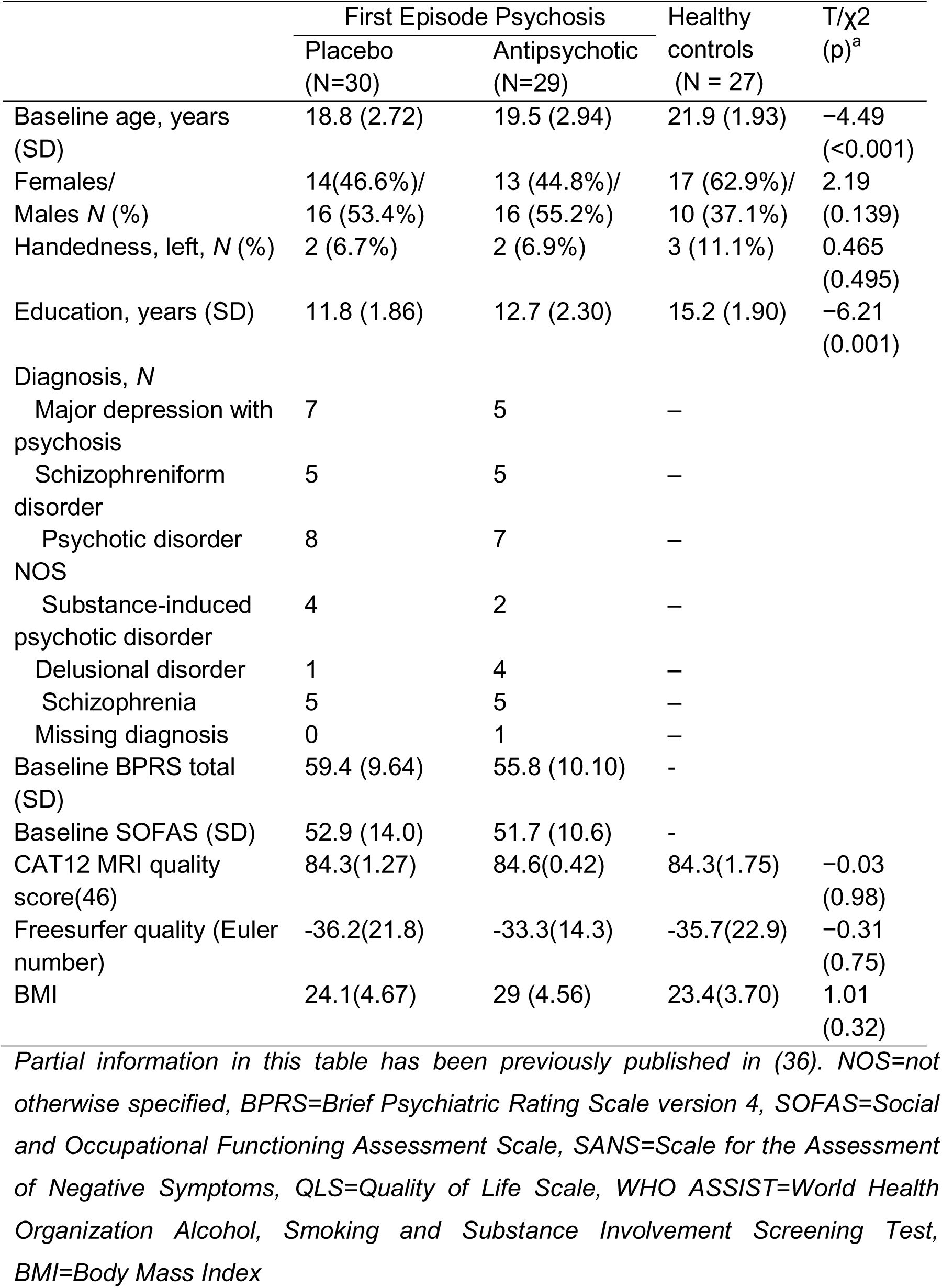
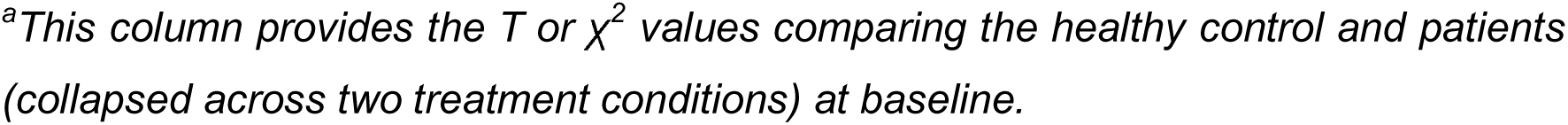
Participant’s demographics.

| | First Episode Psychosis | | Healthy controls | T/$\chi^2$ (p)<sup>a</sup> |
| --- | --- | --- | --- | --- |
|  | Placebo (N=30) | Antipsychotic (N=29) | (N = 27) |  |
| Baseline age, years (SD) | 18.8 (2.72) | 19.5 (2.94) | 21.9 (1.93) | -4.49 (<0.001) |
| Females/ Males N (%) | 14(46.6%)/ 16 (53.4%) | 13 (44.8%)/ 16 (55.2%) | 17 (62.9%)/ 10 (37.1%) | 2.19 (0.139) |
| Handedness, left, N (%) | 2 (6.7%) | 2 (6.9%) | 3 (11.1%) | 0.465 (0.495) |
| Education, years (SD) | 11.8 (1.86) | 12.7 (2.30) | 15.2 (1.90) | -6.21 (0.001) |
| Diagnosis, N |  |  |  |  |
| Major depression with psychosis | 7 | 5 | – |  |
| Schizophreniform disorder | 5 | 5 | – |  |
| Psychotic disorder NOS | 8 | 7 | – |  |
| Substance-induced psychotic disorder | 4 | 2 | – |  |
| Delusional disorder | 1 | 4 | – |  |
| Schizophrenia | 5 | 5 | – |  |
| Missing diagnosis | 0 | 1 | – |  |
| Baseline BPRS total (SD) | 59.4 (9.64) | 55.8 (10.10) | - |  |
| Baseline SOFAS (SD) | 52.9 (14.0) | 51.7 (10.6) | - |  |
| CAT12 MRI quality score(46) | 84.3(1.27) | 84.6(0.42) | 84.3(1.75) | -0.03 (0.98) |
| Freesurfer quality (Euler number) | -36.2(21.8) | -33.3(14.3) | -35.7(22.9) | -0.31 (0.75) |
| BMI | 24.1(4.67) | 29 (4.56) | 23.4(3.70) | 1.01 (0.32) |

*Partial information in this table has been previously published in (36). NOS=not otherwise specified, BPRS=Brief Psychiatric Rating Scale version 4, SOFAS=Social and Occupational Functioning Assessment Scale, SANS=Scale for the Assessment of Negative Symptoms, QLS=Quality of Life Scale, WHO ASSIST=World Health Organization Alcohol, Smoking and Substance Involvement Screening Test, BMI=Body Mass Index*
<sup>a</sup>*This column provides the $T$ or $\chi^2$ values comparing the healthy control and patients (collapsed across two treatment conditions) at baseline.*

### Participants

The treatment group were aged 15–25 years and were people experiencing a first episode of psychosis, defined as fulfilling Structured Clinical Interview for DSM-5 (SCID) criteria for a psychotic disorder, including schizophrenia, schizophreniform disorder, delusional disorder, brief psychotic disorder, major depressive disorder with psychotic symptoms, substance-induced psychotic disorder or psychosis not otherwise specified. Additional inclusion criteria to minimize risk were: ability to provide informed consent; comprehension of English language; no contraindication to MRI scanning; duration of untreated psychosis (DUP) of less than 6 months; living in stable accommodation; low risk to self or others. Two individuals were exposed to antipsychotics at doses lower than the minimal exposure limit for inclusion in our study (<7 days of use or lifetime 1750□mg chlorpromazine equivalent exposure), and the remaining people with psychosis had no prior exposure.

Healthy control participants were aged between 18 and 25, could provide written informed consent, and were psychiatrically, neurologically and medically healthy. Current and historic psychiatric illness was ruled out using self-report, the SCID and The Comprehensive Assessment of At-Risk Mental States (39). In addition, participants were excluded if they were currently taking any psychotropic medications.

A stratified randomization design with sex and DUP as factors was used to allocate people with psychosis to either medication or placebo groups. DUP was included as a three-level factor (0–30, 31–90, and >90 days). Clinicians, patients, study assessors and researchers conducting MRI processing remained blinded to treatment allocation throughout the trial. Further details on inclusion criteria, safety measures, and discontinuation criteria can be found elsewhere (40). A recruitment flow diagram and final group numbers at each timepoint are presented in Supplementary Figure 1, and demographics are presented in Table 1.

### MRI acquisition

A 3-T Siemens Trio Tim scanner located at the Royal Children’s Hospital in Melbourne, Australia, was used to acquire a high-resolution structural T1-weighted Magnetisation-Prepared Rapid Gradient Echo (MPRAGE) scan for each participant. Image acquisition parameters at each timepoint were as follows: 176 sagittal slices, with a 1mm^3^ voxel size, bandwidth 236 Hz/pixel, FOV=256 mm^2^, matrix 256×256, 2300 ms repetition time, and 2.98 ms echo time and a 9° flip angle.

### Brain age models

To estimate each participant’s brain age, two independent, large, previously trained and publicly available models were used. Using these validated pre-trained models enabled robust inference, reproducibility, and cross-study comparison. Both models were trained on very large datasets (>35,000 images) with age ranges that overlap with our sample and have previously reported state-of-the-art performance (41,42). See Table 2 for further details and a comparison between models. Both models have also previously been used to show brain-PAD difference in FEP compared with controls (17,25).

**Table 2.** Summary of the main characteristics of Pyment and CentileBrain models.

|  | **Pymnet** | **CentileBrain** |
| --- | --- | --- |
| **Training sample (n)** | 53,542 | 35,683 |
| **Model** | Simple Fully Convolutional Network | Support Vector Regression with Radial Basis Function kernel |
| **Model Input** | Minimally processed T1 brain volume | 150 morphological features extracted with Freesurfer, Desikan-Killiany atlas (61,62) |
| **Age range** | 3-95 years | 5-90 years |
| **Age bias correction** | No | Yes (63) |
| **Separate age ranges** | No | Yes (5-40 and 41-90 years) |
| **Performance (external dataset)** | MAE (years) = 3.90;<br>r = 0.975 | MAE (years) = 3.56;<br>r = 0.84<sup>a</sup> |
| **Test-retest reliability** | ICC = 0.97 (Ruben P.Dörfel, 2025) | Not yet computed |

MAE = mean absolute error, r = Pearson's correlation coefficient, ICC = interclass correlation coefficient. <sup>a</sup>Reported measurements are not age bias corrected. This is the mean values across the male and female models within a 5-40 years bin.

The primary model we used was Pyment (43), which is a Simple Fully Convolutional Network (SFCN) model that accepts minimally processed T1w MRI images as inputs, after brain extraction using FreeSurfer, rigid registration to the MNI space using FLIRT from FSL (44), and cropping of image borders using nibabel (45) (v. 5.3.2). Pyment does not use age correction (i.e., a correction for the regression towards the mean). To account for possible differences in image quality in our brain age predictions, we additionally used Computational Anatomy Toolbox (version r1113) (46) for the Statistical Parametric Mapping 12 (47) (SPM) to extract Image Quality Rating (IQR) for each raw T1w imaging, to include it as a covariate in all Pyment analyses. Pyment has also been used in longitudinal samples, showing high reliability (48); see Table 2.

The secondary model we used to replicate our findings was CentileBrain (42,49), which uses Support Vector Regression with Radial Basis Function kernel (SVR-RBF). The modelling approach for CentileBrain was originally selected based on performance compared to classical machine learning models. Rather than minimally processed T1w images like Pyment, CentileBrain uses FreeSurfer v7.1 (50) derived grey matter subcortical volume and cortical thickness and surface area estimates. CentileBrain is trained separately for males and females (48), and these models can be accessed using a dedicated web portal created by the authors (https://centilebrain.org). To account for image segmentation quality, we extracted the Euler number for each scan and included it as a covariate in all CentileBrain analyses (52). As both models obtained consistent results, the primary analyses focus on Pyment, with the CentileBrain results detailed in the supplement.

### Statistical analyses

To assess model performance between predicted brain age and chronological age in the control group we calculated the mean absolute error (MAE), weighted MAE (i.e., MAE divided by age range of the sample; wMAE), Pearson correlation coefficient (r) and the proportion of age variance explained by the model (R^2^). To assess test-retest reliability, we calculated the Intraclass Correlation Coefficient (ICC) for brain-PAD across the 3 time points in healthy control participants. The ICC is closer to one when all the values in the time points are similar. As a complementary measure of reliability, we also calculated the Mean Absolute Difference (MAD), which reflects the error between longitudinal scans and equals zero when there is no measurement error.

We used analysis of covariance (ANCOVA) to examine differences in baseline brain-PAD between people with FEP and controls, including age, sex, and a scan quality metric as covariates. White’s (53) method of robust variance estimation was used to account for the non-normality of the residuals. A detailed model assumptions check is provided in Supplementary Table 1. Given that Body Mass Index has been repeatedly found to be related to brain-PAD in schizophrenia samples (26,54), we repeated this analysis after additionally including BMI as a covariate (excluding 8 people [2 controls and 6 people with FEP], with missing BMI data).

To examine differences in longitudinal brain-PAD changes between the three groups (placebo, antipsychotic and healthy controls), we computed a linear mixed-effects model with brain-PAD as outcome and group (healthy control, placebo, antipsychotic), time (in days), age, sex and scan quality measure as fixed effects and subjects as random effects. Our contrast of interest was a group-by-time interaction. A detailed assumptions check for this model is provided in Supplementary Table 2.

Prior to analyses, we compared model fits between a model with only random intercepts and one with both random intercepts and slopes. Models were fitted using maximum likelihood estimation to allow model comparison and a likelihood ratio test was conducted with model comparison assessed using Akaike Information Criterion (AIC) (55). The effect of inter-individual variability was statistically significant for Pyment’s predictions; therefore, a model containing both subject-level random intercepts and slopes was used for the primary analysis. For CentileBrain’s predictions, inter-individual variability of the slopes was negligible, so only the random intercept term was included. Model assumptions and model comparison procedures are reported in the Supplementary Material and Supplementary Tables 2 and 3. For all statistical analyses, variables were z-standardized prior to analysis. All statistical analyses were carried out in R (56) using the ‘lme4’ (57) and ‘lmertest’ (58) packages.

### Exploratory brain-PAD and behavioral associations

We conducted exploratory analyses examining associations between brain-PAD and clinical outcomes, including Social and Occupational Functioning Assessment Scale (SOFAS), the Brief Psychiatric Rating Scale (BPRS-4), Scale for the Assessment of Negative Symptoms (SANS), Quality of Life Scale (QLS) and Hamilton Rating Scales for Depression and Anxiety (HAM-D and HAM-A). We also examined associations with lifestyle factors, including substance use (WHO-ASSIST) and body mass index (BMI). Cross-sectional associations at baseline were assessed using Pearson’s correlation between brain-PAD and each measure, with significance assessed at both corrected (p_FDR_<.05) and uncorrected (p<.05) levels. For longitudinal analyses, we calculated within-participant change scores (12-month follow-up minus baseline) for brain-PAD and each outcome, and tested associations between brain and behavior change scores using Pearson’s correlation.

## Results

### Age prediction

The Pyment model predicted age in healthy controls with low mean absolute error (MAE = 2.26; wMAE = 0.26), although the correlation between observed and predicted age was moderate (r = 0.55; R^2^ = 0.31). This performance is consistent with previous work (42,43) and expected given narrower age ranges typically yielding lower model fits (59). In line with this, studies using similarly restricted age ranges have also reported comparative values (48). Examining model fit using a linear mixed-effect model to account for repeated measurements yielded similar results (R^2^ = 0.27). Figure 2 shows predicted brain age versus chronological age, along with performance metrics for the three groups separately. Both ICC (0.92 [95% CI =0.88 - 0.95]) and MAD (0.95 years) for brain-PAD showed excellent reliability and consistency. Similar results were obtained with the CentileBrain models (see Supplementary Material and Supplementary Figures 2-5).

**Figure 2.**
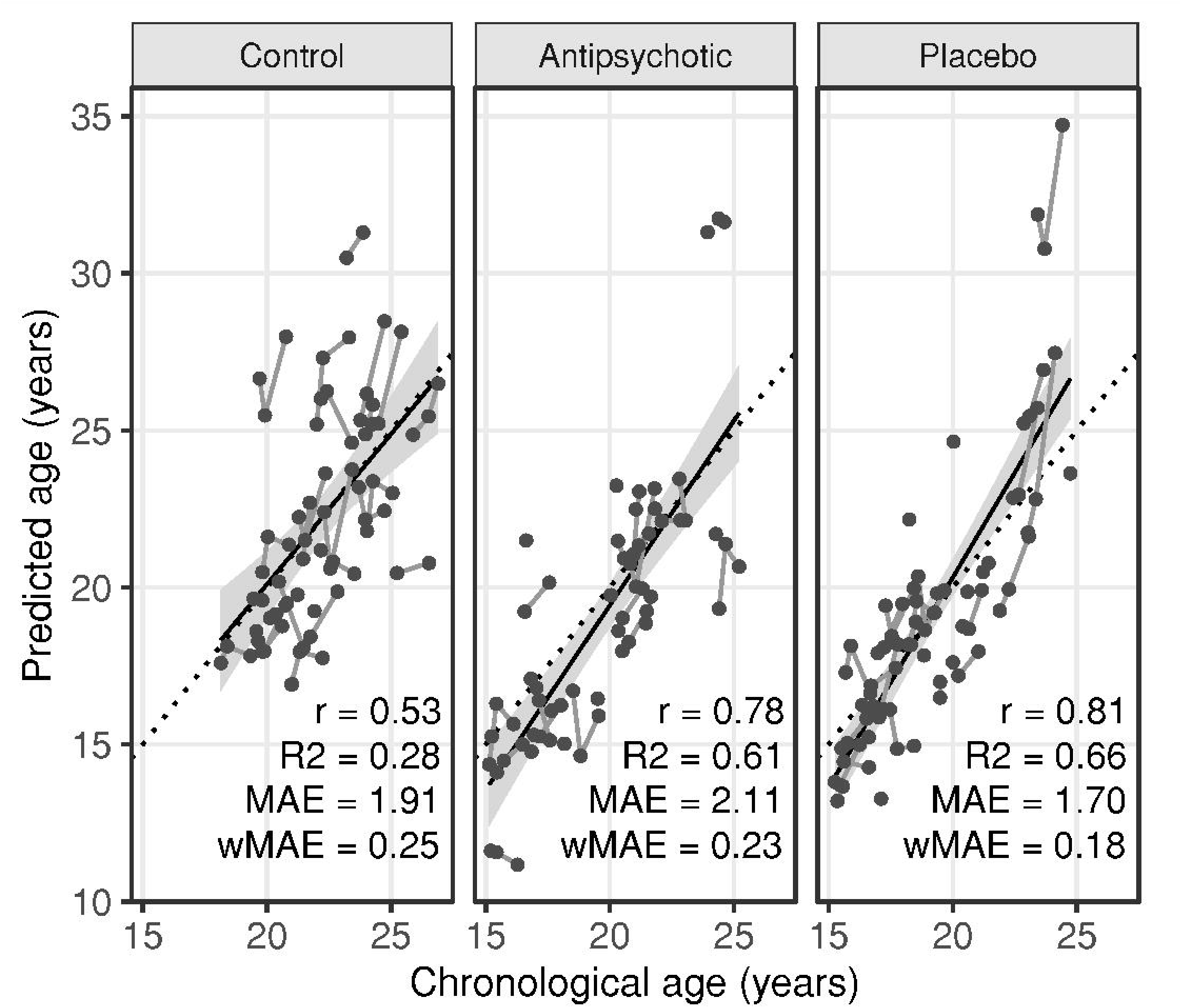
Predicted age versus chronological age. Diagonal dotted lines reflect perfect correlation. Each point corresponds to an individual scan, and each thin line connecting dots represents one individual’s repeated measurements across the study. No covariates are included, as the purpose of this analysis is to evaluate the accuracy of the pre-trained model in predicting age in our sample.

### No difference in baseline brain-PAD in FEP

No significant differences in brain-PAD between FEP group and control group at baseline (F(_1,80_) = 1.30, p = 0.26; Figure 3) were detected. Repeating the analyses after excluding an outlier with extremely high brain-PAD (8.45; SD=3.27) did not alter the results (F(_1,79_) = 0.19, p = 0.94), nor did including BMI as a covariate (F(_1,69_) = 0.80, p = 0.37).

**Figure 3.**
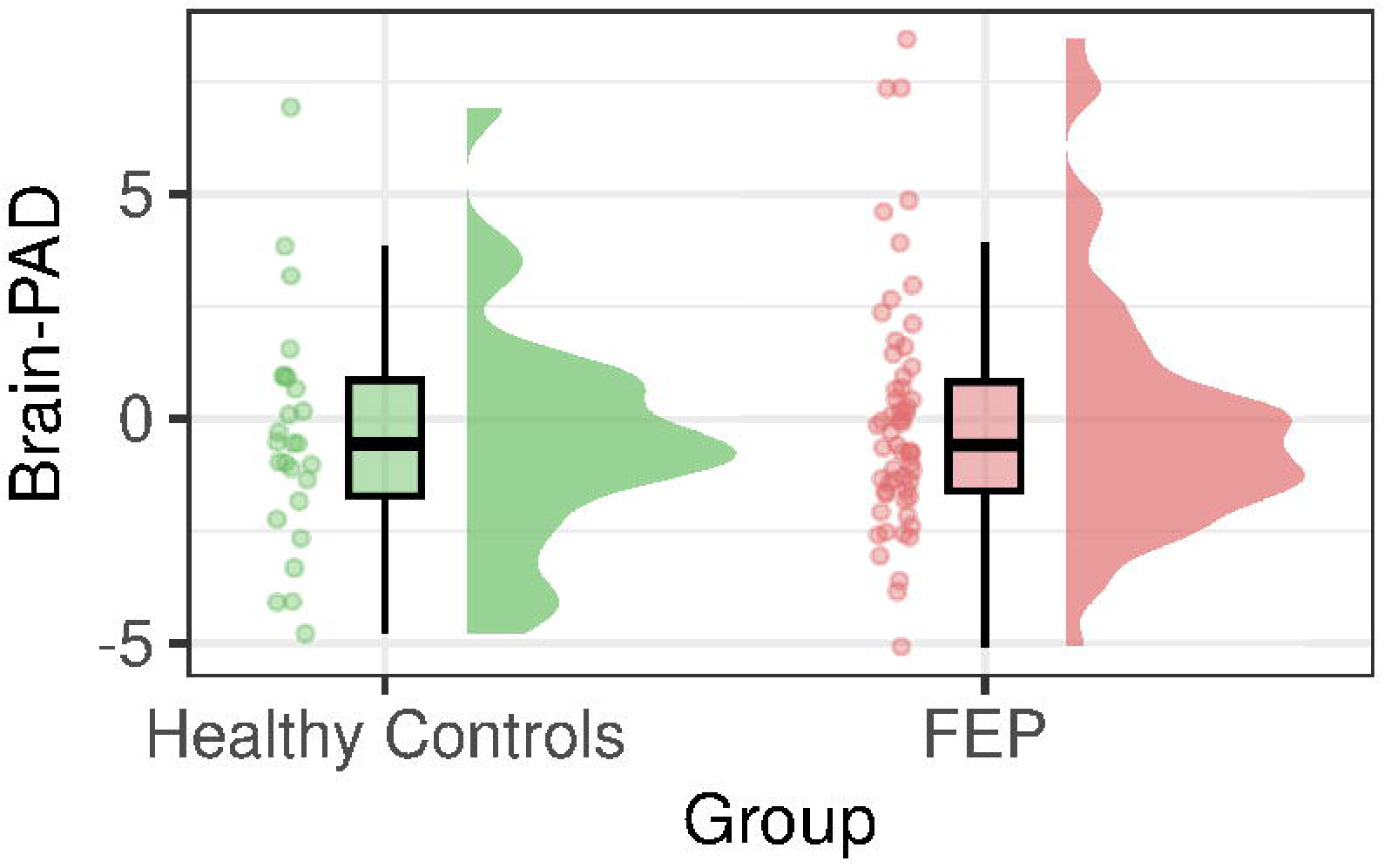
Raincloud plots for brain predicted age difference (Brain-PAD) for healthy controls and First Episode of Psychosis (FEP) groups at baseline. Brain-PAD was predicted using Pyment model and expressed in years. There are no significant differences between the groups.

### Brain-PAD is stable over time and unaffected by antipsychotic medication

There was no statistically significant main effect of time (F_(1,99.13)_ = 0.38, p = 0.54, group (F_(2, 60.54)_ = 0.61, p = 0.55; Figure 4) or interaction between group and time (F_(2, 55.15)_ = 1.38, p = 0.26; Figure 4). Taken together, brain-PAD did not show evidence of change over time, nor any evidence for being impacted by medication exposure or lack thereof.

**Figure 4.**
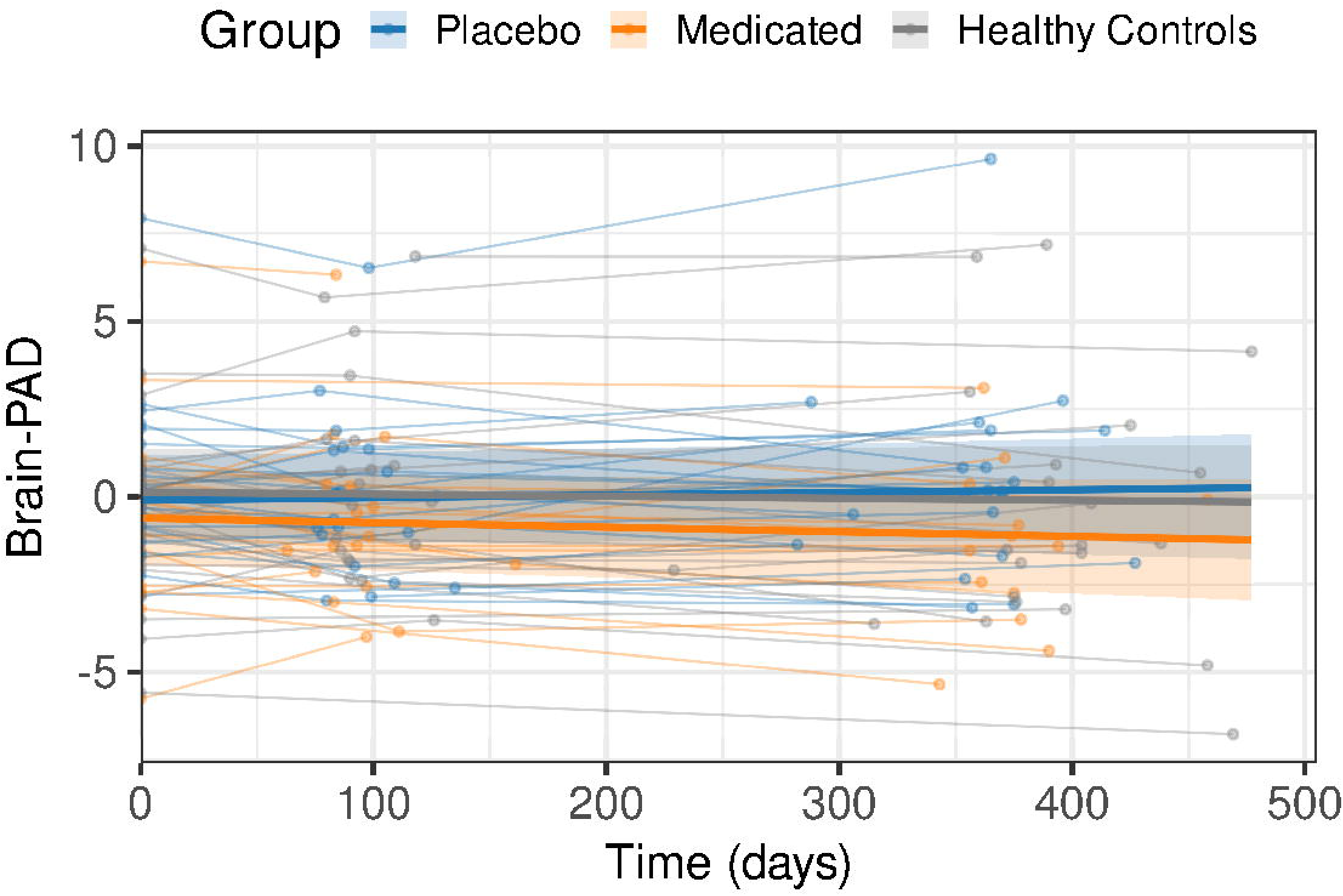
Covariate-adjusted subject-level brain-PAD data with overlaid Linear Mixed Model (LMM) group effects. Dots represent each scan, and dots connected by thin lines represent an individual. LMM group effects are represented in bold lines, where no significant group-by-time interaction was detected. Note that Brain-PAD values are adjusted for age, sex and scan quality.

### Brain-PAD is not associated with clinical variables in FEP

At baseline, no associations were found between clinical variables and brain-PAD (all r< |0.25|; p > 0.14). Similarly, longitudinal change in brain-PAD was not correlated to the change in clinical variables (all r< |0.27|; p > 0.12) at corrected or uncorrected levels.

## Discussion

We used a randomized placebo-controlled design to examine whether antipsychotic-naïve people experiencing a first episode psychosis (FEP) show altered structural brain aging, and whether early antipsychotic medication exposure changes the trajectory of brain aging. We found no differences in brain-predicted age difference (brain-PAD) between FEP and healthy controls at baseline, nor did we observe evidence of any longitudinal differences between patients treated with placebo, antipsychotic medications, or healthy controls over the first year of treatment. Finally, no significant associations were found between brain-PAD, or change in brain-PAD, and clinical measurements. Our findings remained consistent when using two different validated models to estimate brain age.

Our results contrast with those of previous studies in FEP examining brain age reporting differences between patients and healthy controls, despite using the same pretrained brain age prediction models and comparable sample sizes as prior work (17,25). It is possible that differences in sample characteristics, such as symptom severity, affect our results. However, despite strict inclusion criteria to ensure safety (i.e. patients at low risk of harm to themselves or others, living in stable accommodation, and presenting with a short duration of untreated psychosis), the sample of patients used in the current study had similar symptom severity and functioning compared to other published samples (29,33). Other characteristics such as the antipsychotic naïve status and short DUP of our sample would imply that differences in brain ageing may only emerge later during the illness course, as a result of progressive processes, or a cumulation of secondary lifestyle and environmental factors such as substance use, diet and sedentary behaviors.

Our longitudinal findings show that patients who receive antipsychotic medication or placebo show a similar normative brain age trajectory to the healthy control group. These findings are consistent with multiple other studies showing a lack of brain age progression in the early stages of psychosis (29,30,33). Although our findings may appear to diverge from those of Schnack and colleagues (18), who reported progressive brain-PAD accumulation peaking within the first two years of illness, their sample comprised people with schizophrenia on typical antipsychotics, and changes accumulating beyond a one-year window would not be detectable in our study. Taken together, this would suggest that exposure, or lack thereof, to antipsychotics does not alter the stable brain age trajectory in the first year of treatment engagement. One possibility is that the intensive psychosocial treatment provided to both patient groups is sufficient to protect against accelerated brain aging. Additionally, it is possible that only certain patient subgroups exhibit brain aging alterations (30,60). Indeed, one study has reported that brain-PAD alteration in FEP is mainly driven by those who did not show an improvement after risperidone treatment over 3 months (30). In the current trial, participants who failed to show clinical or functional improvement were withdrawn, potentially biasing our sample towards patients who show less accelerated aging. However, as we have previously reported, the clinical and demographic characteristics of those who did and did not complete the final 12-month imaging timepoint did not significantly differ at baseline (36).

While it’s possible that our findings are affected by the performance of the brain age models, the reported values are generally consistent with previously reported performance (43) and are expected to be lower when applied to samples with narrower age ranges (59), such as the current study. In line with this, studies using similarly restricted age ranges have also reported comparative performance (48).

The strengths of the present study include a blinded and placebo-controlled trial design, an antipsychotic-naïve sample, and replication using two large, independent and widely used brain age prediction models. This study had some limitations. The practical challenges of recruiting antipsychotic-naïve individuals limited the sample size of the current cohort, although the size is comparable to prior studies (29,30). Second, our analyses were limited to risperidone and paliperidone, and it therefore remains unclear whether these findings generalize to other antipsychotic medications.

In conclusion, we found no evidence of altered structural brain ageing at illness onset in antipsychotic-naïve FEP, nor of any effect of early antipsychotic exposure on its trajectory over the first year. Brain-age alterations in psychosis may emerge later in later stages, driven by illness progression, cumulative antipsychotic exposure, or secondary lifestyle factors.

## Data Availability

Data is not available

## Ethical Considerations

We certify that formal approval to conduct the experiments described have been obtained from the review board of the Melbourne Health Human Research Ethics Committee (MHREC:2007.616) and could be provided upon request.

## Acknowledgements

This study has been supported by a large number of clinical staff at Orygen Youth Health: Craig Macneil, Kingsley Crisp, Dylan Alexander, Tina Proffitt, Rachel Tindall, Jennifer Hall, Lisa Rumney, Franco Scalzo, Melissa Pane, Linda Kader, Frank Hughes, Clare Shelton, Ryan Kaplan, David Hallford, Bridget Moller, Rick Fraser and research assistants: Daniela Cagliarini, Suzanne Wiltink, Janine Ward and Sumudu Mallawaarachichi. The trial took place at the Early Psychosis Prevention and Intervention Centre, which is part of Orygen Youth Health, Melbourne, Australia. Alejandro Roig Herrer was supported by research grants PRE2022-104038 funded by MCIN/AEI/10.13039/501100011033 and the ESF+. Barnaby Nelson was supported by an NHMRC Senior Research Fellowship (1137687). Christos Pantelis was supported by a National Health and Medical Research Council (NHMRC) L3 Investigator Grant (1196508). Laura Han was supported by the NWO-VENI grant (number 09150162210201). Sidhant Chopra was supported by the University of Melbourne McKenzie Fellowship and Brain Behaviour Foundation Young Investigator Grant. This work was also funded by MCIN/AEI/10.13039/501100011033 through grants PID2021-124407NB-I00 and PID2024-162453NB-I00, co-funded by the European Union, European Regional Development Fund (ERDF, 2021–2027).

## References

1. Shivakumar V, Kalmady S V, Venkatasubramanian G, Ravi V, Gangadhar BN (2014): Do schizophrenia patients age early? Asian J Psychiatr 10: 3–9.

2. Blose BA, Lai A, Crosta C, Thompson JL, Silverstein SM (2023): Retinal Neurodegeneration as a Potential Biomarker of Accelerated Aging in Schizophrenia Spectrum Disorders. Schizophr Bull 49: 1316–1324.

3. Tian YE, Cropley V, Maier AB, Lautenschlager NT, Breakspear M, Zalesky A (2023): Heterogeneous aging across multiple organ systems and prediction of chronic disease and mortality. Nat Med 29: 1221–1231.

4. Zheng F, Liu Y, Yuan Z, Gao X, He Y, Liu X, et al. (2019): Age□related changes in cortical and subcortical structures of healthy adult brains: A surface□based morphometry study. Journal of Magnetic Resonance Imaging 49: 152–163.

5. Franke K, Ziegler G, Klöppel S, Gaser C, Initiative ADN (2010): Estimating the age of healthy subjects from T1-weighted MRI scans using kernel methods: exploring the influence of various parameters. Neuroimage 50: 883–892.

6. Blake K V., Ntwatwa Z, Kaufmann T, Stein DJ, Ipser JC, Groenewold NA (2023): Advanced brain ageing in adult psychopathology: A systematic review and meta-analysis of structural MRI studies. J Psychiatr Res 157: 180–191.

7. Ballester PL, Romano MT, de Azevedo Cardoso T, Hassel S, Strother SC, Kennedy SH, Frey BN (2022): Brain age in mood and psychotic disorders: a systematic review and meta-analysis. Acta Psychiatr Scand 145: 42–55.

8. Wrigglesworth J, Ward P, Harding IH, Nilaweera D, Wu Z, Woods RL, Ryan J (2021): Factors associated with brain ageing - a systematic review. BMC Neurol 21: 312.

9. Chen C, Hwang T, Tung Y, Yang L, Hsu Y, Liu C, et al. (2022): Detection of advanced brain aging in schizophrenia and its structural underpinning by using normative brain age metrics. Neuroimage Clin 34: 103003.

10. Constantinides C, Han LKM, Alloza C, Antonucci LA, Arango C, Ayesa-Arriola R, et al. (2023): Brain ageing in schizophrenia: evidence from 26 international cohorts via the ENIGMA Schizophrenia consortium. Mol Psychiatry 28: 1201–1209.

11. Kaufmann T, van der Meer D, Doan NT, Schwarz E, Lund MJ, Agartz I, et al. (2019): Common brain disorders are associated with heritable patterns of apparent aging of the brain. Nat Neurosci 22: 1617–1623.

12. Kim WS, Heo DW, Maeng J, Shen J, Tsogt U, Odkhuu S, et al. (2024): Deep Learning-based Brain Age Prediction in Patients With Schizophrenia Spectrum Disorders. Schizophr Bull 50: 804–814.

13. Kim WS, Heo DW, Shen J, Tsogt U, Odkhuu S, Kim SW, et al. (2023): Stage-Specific Brain Aging in First-Episode Schizophrenia and Treatment-Resistant Schizophrenia. International Journal of Neuropsychopharmacology 26: 207–216.

14. Kuo C-Y, Lee P-L, Hung S-C, Liu L-K, Lee W-J, Chung C-P, et al. (2020): Large-Scale Structural Covariance Networks Predict Age in Middle-to-Late Adulthood: A Novel Brain Aging Biomarker. Cerebral Cortex 30: 5844–5862.

15. Lieslehto J, Jääskeläinen E, Kiviniemi V, Haapea M, Jones PB, Murray GK, et al. (2021): The progression of disorder-specific brain pattern expression in schizophrenia over 9 years. NPJ Schizophr. 10.1038/s41537-021-00157-0

16. Nenadić I, Dietzek M, Langbein K, Sauer H, Gaser C (2017): BrainAGE score indicates accelerated brain aging in schizophrenia, but not bipolar disorder. Psychiatry Res Neuroimaging 266: 86–89.

17. Roig-Herrero A, San-José-Revuelta LM, Navarro-González R, de Luis-García R, Molina V (2026): Effects of disease duration and antipsychotics on brain age in schizophrenia. Schizophr Res 287: 82–90.

18. Schnack HG, Van Haren NEM, Nieuwenhuis M, Pol HEH, Cahn W, Kahn RS (2016): Accelerated brain aging in schizophrenia: A longitudinal pattern recognition study. American Journal of Psychiatry 173: 607–616.

19. Shahab S, Mulsant BH, Levesque ML, Calarco N, Nazeri A, Wheeler AL, et al. (2019): Brain structure, cognition, and brain age in schizophrenia, bipolar disorder, and healthy controls. Neuropsychopharmacology 44: 898–906.

20. Zhu JD, Tsai SJ, Lin CP, Lee YJ, Yang AC (2023): Predicting aging trajectories of decline in brain volume, cortical thickness and fractional anisotropy in schizophrenia. Schizophrenia 9. 10.1038/s41537-022-00325-w

21. Zhu J-D, Wu Y-F, Tsai S-J, Lin C-P, Yang AC (2023): Investigating brain aging trajectory deviations in different brain regions of individuals with schizophrenia using multimodal magnetic resonance imaging and brain-age prediction: a multicenter study. Transl Psychiatry 13: 82.

22. Kolenic M, Franke K, Hlinka J, Matejka M, Capkova J, Pausova Z, et al. (2018): Obesity, dyslipidemia and brain age in first-episode psychosis. J Psychiatr Res 99: 151–158.

23. Bittner N, Jockwitz C, Franke K, Gaser C, Moebus S, Bayen UJ, et al. (2021): When your brain looks older than expected: combined lifestyle risk and BrainAGE. Brain Struct Funct 226: 621–645.

24. Andreasen NC, Liu D, Ziebell S, Vora A, Ho B-C (2013): Relapse duration, treatment intensity, and brain tissue loss in schizophrenia: a prospective longitudinal MRI study. American Journal of Psychiatry 170: 609–615.

25. Hua JPY, Abram S V, Loewy RL, Stuart B, Fryer SL, Vinogradov S, Mathalon DH (2024): Brain Age Gap in Early Illness Schizophrenia and the Clinical High-Risk Syndrome: Associations With Experiential Negative Symptoms and Conversion to Psychosis. Schizophr Bull 50: 1159–1170.

26. Kolenič M, McWhinney SR, Selitser M, Šafářová N, Franke K, Vochoskova K, et al. (2025): Central Obesity-related Brain Alterations Predict Cognitive Impairments in First Episode of Psychosis. Schizophr Bull sbaf101.

27. Hajek T, Franke K, Kolenic M, Capkova J, Matejka M, Propper L, et al. (2019): Brain Age in Early Stages of Bipolar Disorders or Schizophrenia. Schizophr Bull 45: 191–198.

28. Truelove-Hill M, Erus G, Bashyam V, Varol E, Sako C, Gur RC, et al. (2020): A Multidimensional Neural Maturation Index Reveals Reproducible Developmental Patterns in Children and Adolescents. The Journal of Neuroscience 40: 1265.

29. Yi-Bin X, Xu-Sha W, Cui L-B, Li-Jun B, Gan S-Q, Xiao-Yan J, et al. (2022): Neuroimaging-based brain-age prediction of first-episode schizophrenia and the alteration of brain age after early medication. The British Journal of Psychiatry 220: 339–346.

30. Fan L, Zhang Z, Ma X, Liang L, Yuan L, Ouyang L, et al. (2025): Brain Age Gap as a Predictor of Early Treatment Response and Functional Outcomes in First-Episode Schizophrenia: A Longitudinal Study: L’écart d’âge cérébral comme prédicteur de la réponse en début de traitement et des résultats fonctionnels dans un prem. Can J Psychiatry 70: 240–250.

31. Chung Y, Addington J, Bearden CE, Cadenhead K, Cornblatt B, Mathalon DH, et al. (2018): Use of Machine Learning to Determine Deviance in Neuroanatomical Maturity Associated With Future Psychosis in Youths at Clinically High Risk. JAMA Psychiatry 75: 960–968.

32. Salisbury DF, Wulf BM, Seebold D, Coffman BA, Curtis MT, Karim HT (2024): Predicted Brain Age in First-Episode Psychosis: Association with Inexpressivity. Brain Sciences, vol. 14. 10.3390/brainsci14060532

33. Mcwhinney S, Kolenic M, Franke K, Fialova M, Knytl P, Matejka M, et al. (2021): Obesity as a Risk Factor for Accelerated Brain Ageing in First-Episode Psychosis-A Longitudinal Study. Schizophr Bull 47: 1772–1781.

34. Zhu J-D, Tsai S-J, Lin C-P, Lee Y-J, Yang AC (2023): Predicting aging trajectories of decline in brain volume, cortical thickness and fractional anisotropy in schizophrenia. Schizophrenia 9: 1.

35. Francey SM, O’Donoghue B, Nelson B, Graham J, Baldwin L, Yuen HP, et al. (2020): Psychosocial Intervention With or Without Antipsychotic Medication for First-Episode Psychosis: A Randomized Noninferiority Clinical Trial. Schizophr Bull Open 1: sgaa015.

36. Chopra S, Fornito A, Francey SM, O’Donoghue B, Cropley V, Nelson B, et al. (2021): Differentiating the effect of antipsychotic medication and illness on brain volume reductions in first-episode psychosis: A Longitudinal, Randomised, Triple-blind, Placebo-controlled MRI Study. Neuropsychopharmacology 46: 1494–1501.

37. Chopra S, Holmes A, Segal A, Zhang X-H, Francey SM, O’Donoghue B, et al. (2025): Differential effects of illness and antipsychotics on cortical thinning in first episode psychosis: A randomised placebo-controlled MRI study. medRxiv 2025.

38. Francey SM, O’Donoghue B, Nelson B, Graham J, Baldwin L, Yuen HP, et al. (2020): Psychosocial Intervention With or Without Antipsychotic Medication for First-Episode Psychosis: A Randomized Noninferiority Clinical Trial. Schizophr Bull Open 1: sgaa015.

39. Yung AR, Yung AR, Pan Yuen H, Mcgorry PD, Phillips LJ, Kelly D, et al. (2005): Mapping the onset of psychosis: the comprehensive assessment of at-risk mental states. Australian and New Zealand Journal of Psychiatry 39: 964–971.

40. O’Donoghue B, Francey SM, Nelson B, Ratheesh A, Allott K, Graham J, et al. (2019): Staged treatment and acceptability guidelines in early psychosis study (STAGES): A randomized placebo controlled trial of intensive psychosocial treatment plus or minus antipsychotic medication for first□episode psychosis with low□risk of self□harm or aggression. Study protocol and baseline characteristics of participants. Early Interv Psychiatry 13: 953–960.

41. Gaser C, Dahnke R, Thompson PM, Kurth F, Luders E, Initiative ADN (2024): CAT: a computational anatomy toolbox for the analysis of structural MRI data. Gigascience 13: giae049.

42. Dörfel RP, Arenas□Gomez JM, Fisher PM, Ganz M, Knudsen GM, Svensson JE, Plavén□Sigray P (2023): Prediction of brain age using structural magnetic resonance imaging: A comparison of accuracy and test–retest reliability of publicly available software packages. Hum Brain Mapp 44: 6139–6148.

43. Yu Y, Cui H, Haas SS, New F, Sanford N, Yu K, et al. (2024): Brain□age prediction: Systematic evaluation of site effects, and sample age range and size. Hum Brain Mapp 45: e26768.

44. Desikan RS, Ségonne F, Fischl B, Quinn BT, Dickerson BC, Blacker D, et al. (2006): An automated labeling system for subdividing the human cerebral cortex on MRI scans into gyral based regions of interest. Neuroimage 31: 968–980.

45. Klein A, Tourville J (2012): 101 labeled brain images and a consistent human cortical labeling protocol. Front Neurosci 6: 33392.

46. Beheshti I, Nugent S, Potvin O, Duchesne S (2019): Bias-adjustment in neuroimaging-based brain age frameworks: A robust scheme. Neuroimage Clin 24: 102063.

47. Leonardsen EH, Peng H, Kaufmann T, Agartz I, Andreassen OA, Celius EG, et al. (2022): Deep neural networks learn general and clinically relevant representations of the ageing brain. Neuroimage 256: 119210.

48. Jenkinson M, Bannister P, Brady M, Smith S (2002): Improved optimization for the robust and accurate linear registration and motion correction of brain images. Neuroimage 17: 825–841.

49. Brett M, Markiewicz CJ, Hanke M, Côté M-A, Cipollini B, McCarthy P, et al. (2020): nipy/nibabel. Zenodo.

50. Tzourio-Mazoyer N, Landeau B, Papathanassiou D, Crivello F, Etard O, Delcroix N, et al. (2002): Automated anatomical labeling of activations in SPM using a macroscopic anatomical parcellation of the MNI MRI single-subject brain. Neuroimage 15: 273–289.

51. Han LKM, Dehestani N, Suo C, Daglas-Georgiou R, Hasty M, Kader L, et al. (2025): Longitudinal brain age in first-episode mania youth treated with lithium or quetiapine. European Neuropsychopharmacology 95: 40–48.

52. Ge R, Yu Y, Qi YX, Fan Y, Chen S, Gao C, et al. (2024): Normative modelling of brain morphometry across the lifespan with CentileBrain: algorithm benchmarking and model optimisation. Lancet Digit Health 6: e211–e221.

53. Fischl B (2012): FreeSurfer. Neuroimage 62: 774–781.

54. Sanford N, Ge R, Antoniades M, Modabbernia A, Haas SS, Whalley HC, et al. (2022): Sex differences in predictors and regional patterns of brain age gap estimates. Hum Brain Mapp 43: 4689–4698.

55. Rosen AFG, Roalf DR, Ruparel K, Blake J, Seelaus K, Villa LP, et al. (2018): Quantitative assessment of structural image quality. Neuroimage 169: 407–418.

56. White H (1980): A Heteroskedasticity-Consistent Covariance Matrix Estimator and a Direct Test for Heteroskedasticity. Econometrica 48: 817–838.

57. Ronan L, Alexander-Bloch AF, Wagstyl K, Farooqi S, Brayne C, Tyler LK, Fletcher PC (2016): Obesity associated with increased brain age from midlife. Neurobiol Aging 47: 63–70.

58. Pinheiro JC, Bates DM (2000): Theory and computational methods for linear mixed-effects models. Mixed-effects models in S and S-PLUS 57–96.

59. Team RC (2021): R: A language and environment for statistical computing. R foundation for statistical computing, Vienna, Austria.

60. Bates D, Maechler M, Bolker B, Walker S, Christensen RHB, Singmann H, et al. (2015): Package ‘lme4.’ convergence 12: 2.

61. Kuznetsova A, Brockhoff PB, Christensen RHB (2015): Package ‘lmertest.’ R package version 2: 734.

62. de Lange AG, Anatürk M, Rokicki J, Han LKM, Franke K, Alnæs D, et al. (2022): Mind the gap: Performance metric evaluation in brain□age prediction. Hum Brain Mapp 43: 3113–3129.

63. Haas SS, Ge R, Sanford N, Modabbernia A, Reichenberg A, Whalley HC, et al. (2022): Accelerated Global and Local Brain Aging Differentiate Cognitively Impaired From Cognitively Spared Patients With Schizophrenia. Front Psychiatry 13. 10.3389/fpsyt.2022.913470

