## Supplementary for "Structural Brain Age Trajectories in Antipsychotic-naive First Episode Psychosis"

**Contents:**

Supplementary Table 1 – Cumulative antipsychotic exposure

Supplementary Figure 1: The flow of patients and healthy controls through the study

Supplementary table 2 – Summary of the main characteristics of Pyment and CentileBrain models

Supplementary table 3 – Assumptions of the baseline ANCOVAs

Supplementary table 4 – Assumptions of the mixed effect linear models

Supplementary table 5 – Model fit comparison between random intercepts or random intercepts and slopes.

**Results using the CentileBrain model for age prediction**

Supplementary Figure 2. Predicted age versus chronological age.

Supplementary Figure 3: Boxplots for brain predicted age difference (Brain-PAD)	for each of the groups in baseline.

Supplementary Figure 4. Adjusted subject-level data with overlaid Linear Mixed	Model (LMM) group effects.

Supplementary figure 5. Longitudinal association between olanzapine exposure	and brain aging.

**Supplementary material**

Supplementary Table 1 – Cumulative antipsychotic exposure (in Olanzapine milligram equivalates). This table was previously published in Chopra et al 2021.

|  | Baseline | 3 months | 12 months |
| --- | --- | --- | --- |
| PIPT, mg (M, SD) | 0.16 (0.59) | 78.5 (216)* | 608 (1111) |
| MIPT, mg (M, SD) | 1.03 (5.57) | 420 (248) | 1311 (1011) |

*Note: In our primary analysis, the 4 patients within the placebo (PIPT) group who were exposed to antipsychotic medication at amounts greater than the study inclusion criteria at the 3-month timepoint were excluded from the analysis, thus cumulative antipsychotic exposure of the analysis sample was 4.57mg (14.5)

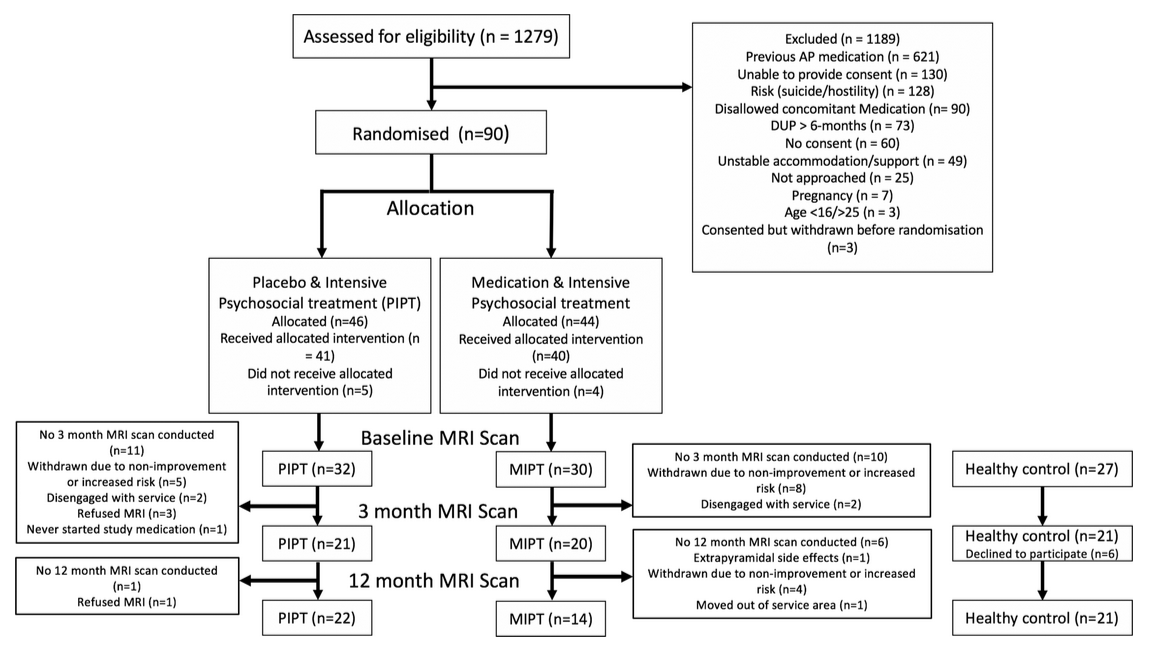
Supplementary Figure 1: The flow of patients and healthy controls through the study. This figure was previously published in Chopra et al 2021.

**Material and methods**

Supplementary table 2 – Summary of the main characteristics of Pyment and CentileBrain models.

|  | Pyment | CentileBrain |
| --- | --- | --- |
| Training set sample | 53542 | 35683 |
| Model | Simple Fully Convolutional Network | Support Vector Regresion with Radial Basis Function kernel |
| Model Input | Barely processed T1 brain volume | 150 morphological features extracted with Freesurfer |
| Age range | 3-95 years | 5-90 years |
| Age bias correction | No | Yes (Betshedi et al 2019) |
| Separe age ranges | No | Yes (5-40 and 41-90 years) |
| Performance (external dataset) | MAE = 3.90, r =0.975 | MAE = 3.56, r = 0.84^a^ |
| Follow-up consistency | ICC = (0.97) (Ruben P.Dörfel, 2025) | Not yet computed |

MAE mean absolute error, r Pearson’s correlation coefficient, ICC interclass correlation coefficient. ^a^Reported measurements are not age bias corrected. It is the mean values of males and females models within 5-40 years bin.

Supplementary table 3 – Assumptions of the baseline ANCOVAs

|  | Pyment | CentileBrain |
| --- | --- | --- |
| Normality of the residuals | Shapiro-Wilk test, W = 0.93, p =0.0003 | Shapiro-Wilk test, W = 0.98, p =0.10 |
| Homocedasticity of the residuals | Breusch-Pagan test: 0.84, p =0.93 | Breusch-Pagan test: 8.7, p =0.07 |
| Mean 0 of the residuals of each group | Controls: *p*-value = 1 | Controls: *p*-value = 1 |
|  | Patients: *p*-value = 1 | Patients: *p*-value = 1 |
| Homogeneity of within slopes | Interactive models do not provide more variance explanation in comparison to the parsimonious ones. | |
| Fixed grouping variable | The grouping variable is fixed because it represents predefined, distinct categories. | |
| Independance of observations | All observations are independent (i.e. no repeated measures). | |

Supplementary Table 4 shows a slight violation of heteroscedasticity and non-normality of the residuals for the Pyment and the CentileBrain model respectively. Nevertheless, violations of these assumptions are proved not to be relevant, particularly when there is no combination of violations, for further information check *Robustness of linear mixed-effects models to violations of distributional assumptions* of Schielzeth et al, 2020.

Supplementary table 4 – Assumptions of the mixed effect linear models

|  | Pyment | CentileBrain |
| --- | --- | --- |
| Normality of the residuals | Shapiro-Wilk test, W = 0.99, p =0.24 | Shapiro-Wilk test, W = 0.98, p =0.007 |
| Homocedasticity of the residuals | Breusch-Pagan test: 4.9, p =0.03 | Breusch-Pagan test: 8.7, p =0.07 |
| Mean 0 of the residuals of each group | Placebo: *p*-value = 0.98 | Placebo: *p*-value = 0.88 |
|  | Medicated: *p*-value = 0.95 | Medicated: *p*-value = 0.88 |
|  | Controls: p-value = 0.98 | Controls: p-value = 0.69 |
| Homogeneity of within slopes | Interactive models do not provide more variance explanation in comparison to the parsimonious ones. | |
| Fixed grouping variable | The grouping variable is fixed because it represents predefined, distinct categories. | |

Model characterisation was compared to determine the appropriate random effect terms for the mixed effects linear models. To test whether individual variability in longitudinal change warranted a random-slope structure, we compared two linear mixed-effects models: one with a random intercept for subject and another with a random intercept and a random slope for time. Models were fitted using maximum likelihood estimation (ML), and a likelihood ratio test was conducted. As seen in Supplementary Table 5, the reduction off the Akaike Information Criterion (AIC) implies that random slopes and intercepts is a better fit for Pyment’s predictions, further assessed by the likelihood ratio test (LRT). In contrast, CentileBrain does not benefit from the inclusion of random slopes, as indicated by an increase in AIC and non-significant LRT.

Supplementary table 5 – Model fit comparison between random intercepts or random intercepts and slopes.

| **Pyment** | | | | |
| --- | --- | --- | --- | --- |
| Model | AIC | logLIK | ΔAIC | p-value (LRT) |
| Random intecept | 682.36 | -330.18 | - | - |
| Random slopes and intercepts | 674.20 | -324.10 | -8.16 | 0.0023 |
| **CentileBrain** | | | | |
| Random intecept | 747.94 | -362.97 | - | - |
| Random slopes and intercepts | 751.81 | -362.90 | +3.87 | 0.93 |

AIC Akaike Information Criterion, logLIK log-likelihood, LRT likelihood ratio test.

**Results using the CentileBrain model for age prediction:**

A) **Age prediction:** Model performance was evaluated in the healthy control group. The model predicted young healthy controls in baseline with low absolute error, MAE = 2.26 (weighted MAE = 0.29) although its correlation was moderate, r = 0.45, R^2^ = 0.20. Fitting a linear mixed effect model to take repeated measurements into account does not vary substantially its fit (R2 = 0.16). Supplementary Figure 2 shows the predicted brain age against chronological age, along with performance metrics in baseline, for the three groups.

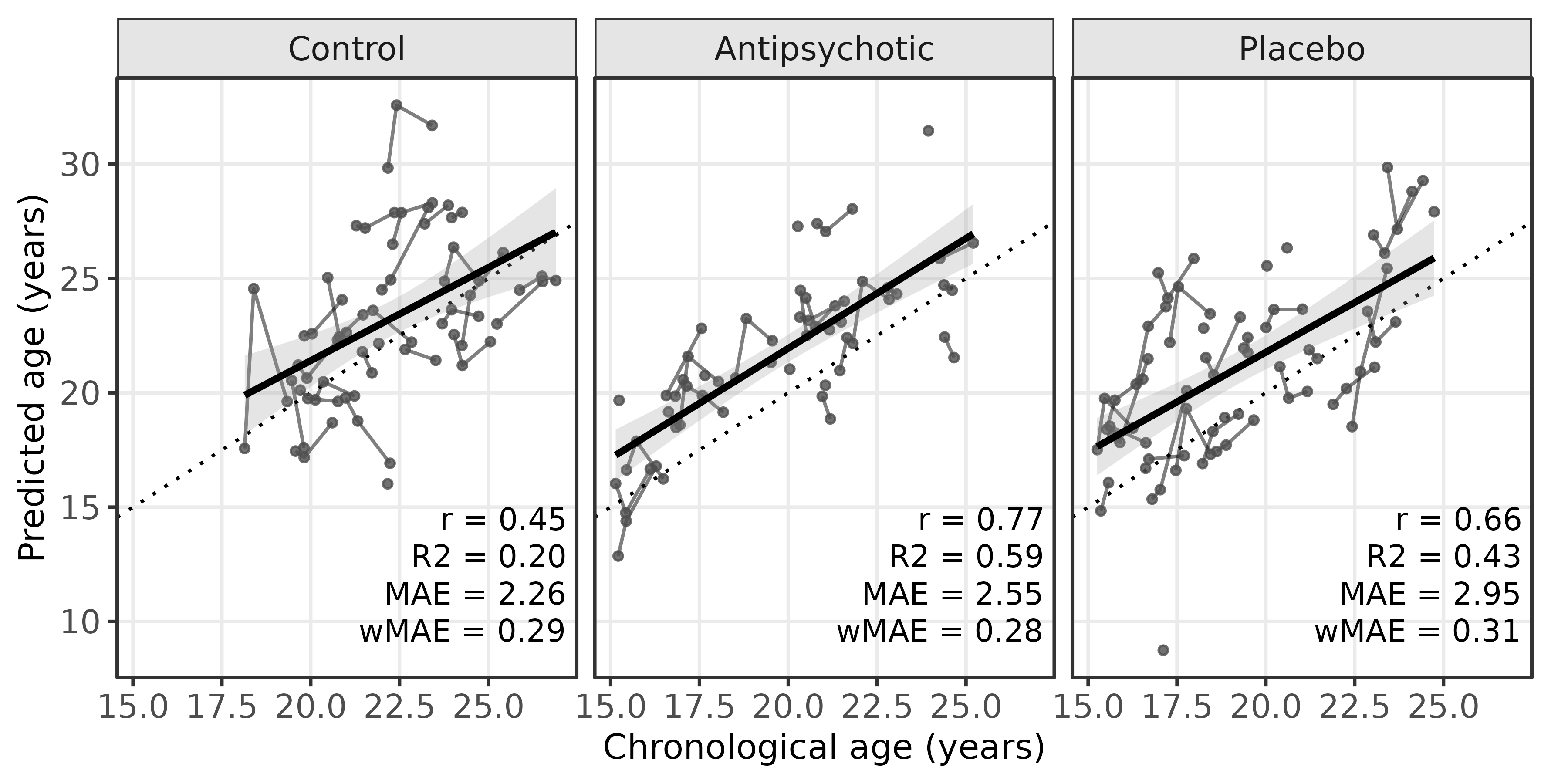
Suplpementary Figure 2. Predicted age versus chronological age. Diagonal dotted lines reflect perfect correlation. Each dot corresponds to an individual scan while each line connecting dots represents one individual's repeated measurements across the study. As intended to represent the accuracy of the predictive model, regression lines are not longitudinally modeled, and no covariates are included.

B) **Brain-PAD is reliable and accurate over time:** Both ICC and MAD showed high reliability and consistency. The ICC for the brain PAD was 0.81 [0.71 - 0.88] and the MAD was 1.35 years.
C) **No difference in baseline brain-PAD in FE patients:** ANCOVA results showed no significant differences in Brain-PAD between FEP patients and controls at baseline, F_(1,80)_ = 1.41, p = 0.23 (Supplementary Figure 3). Repeating the analyses after excluding an outlier with extremely low brain-PAD did not alter the results, nor did including BMI as a covariate F_(1,69)_ = 1.51, p = 0.22.

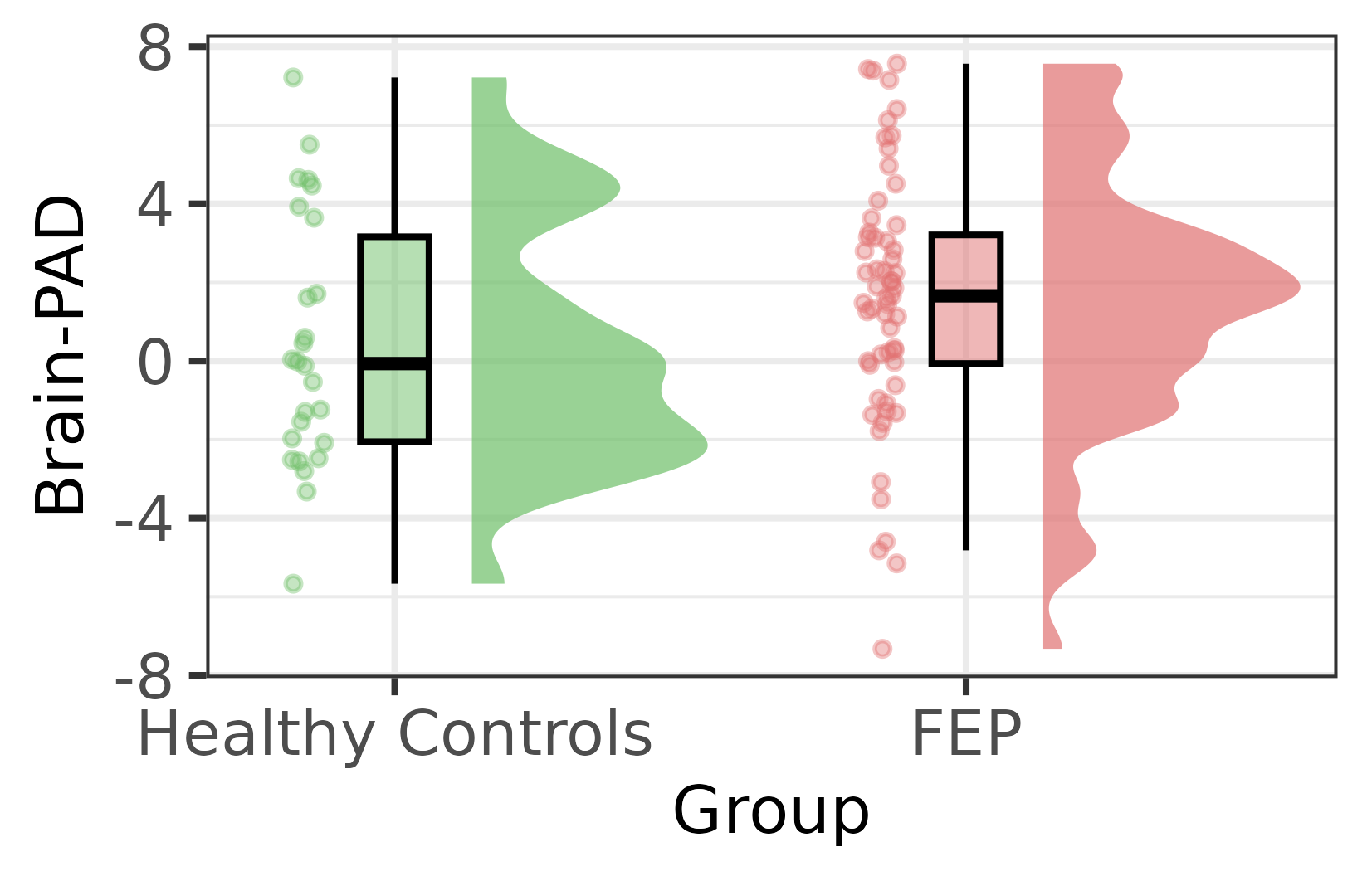
Supplementary Figure 3: Boxplots for brain predicted age difference (Brain-PAD) for each of the groups in baseline, healthy controls and First Episode of Psychosis (FEP). Brain-PAD is expressed in years. There are no significant differences between the groups.

D) **No change in brain-PAD over time for patients:** There was no statistically significant effect of time (F_(1,163.51)_ = 0.41, p = 0.52), group (F_(2, 60.85)_ = 0.04, p = 0.95) or interaction between group and time (F_(2, 109.77)_ = 2.24, p = 0.11). Taken together, brain-PAD did not show evidence of change over time, nor any evidence for differences between the groups over time. Individual and group trajectories of brain-PAD can be seen in Supplementary Figure 4.
E) **Brain-PAD is not associated with symptoms severity:** At baseline, no associations were found between measures of symptoms and brain-PAD (all r< |0.24|; p > 0.07). When the longitudinal change of brain-PAD was correlated to the change in other variables, similar results were found (all r< |0.26|; p > 0.14) except for the change in brain-PAD versus change in the cumulative dose of olanzapine. A post hoc descriptive analysis of this correlation showed that the correlation was highly influenced by an outlier (see Supplementary Figure 5).

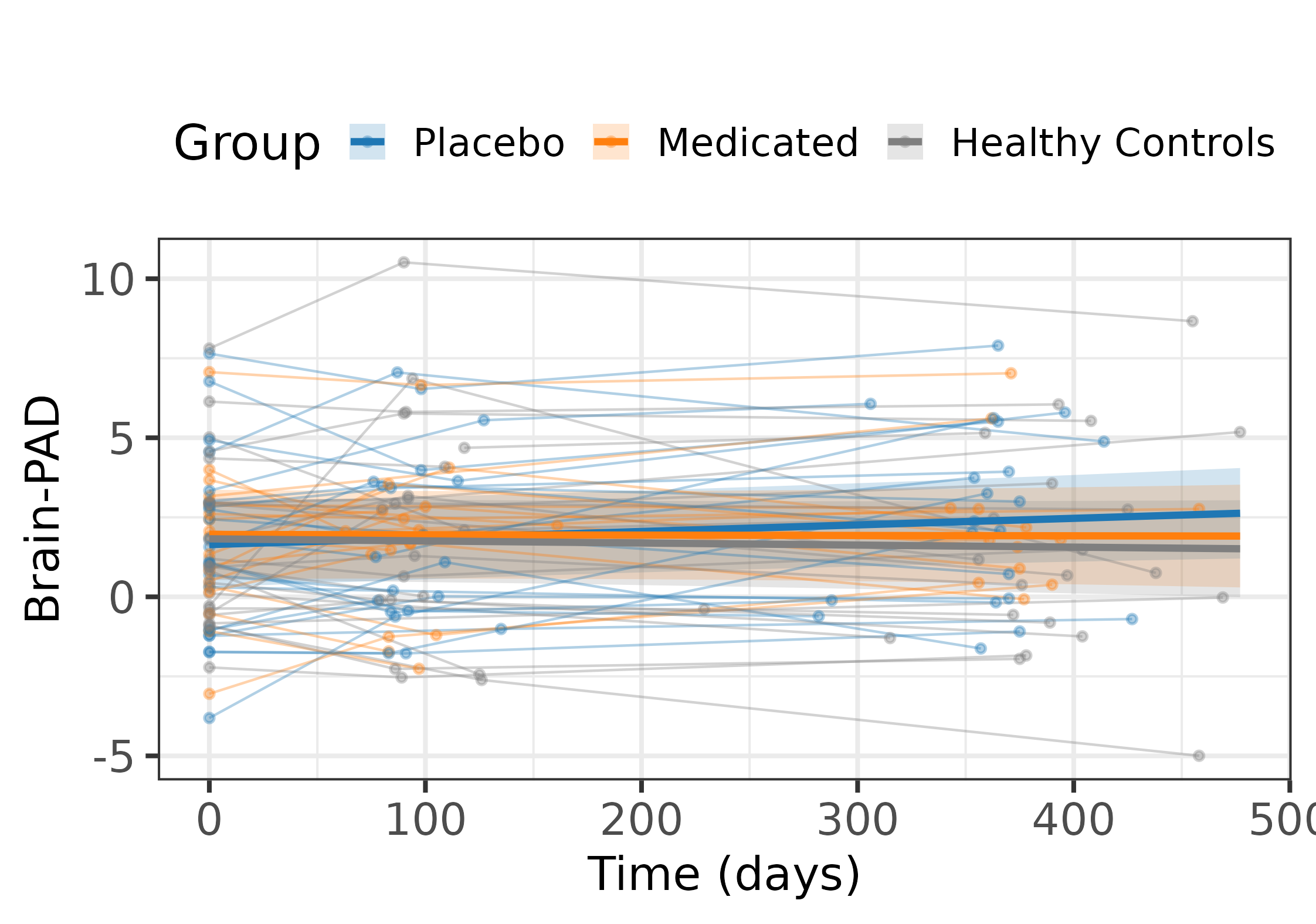
Supplementary Figure 4. Adjusted subject-level data with overlaid Linear Mixed Model (LMM) group effects. Dots represent each scan, and dots connected by thin lines represent an individual. LMM group effects are represented in bold lines, where no significant group-by-time interaction was detected. Note that Brain-PAD values are adjusted for age, sex and Euler number.

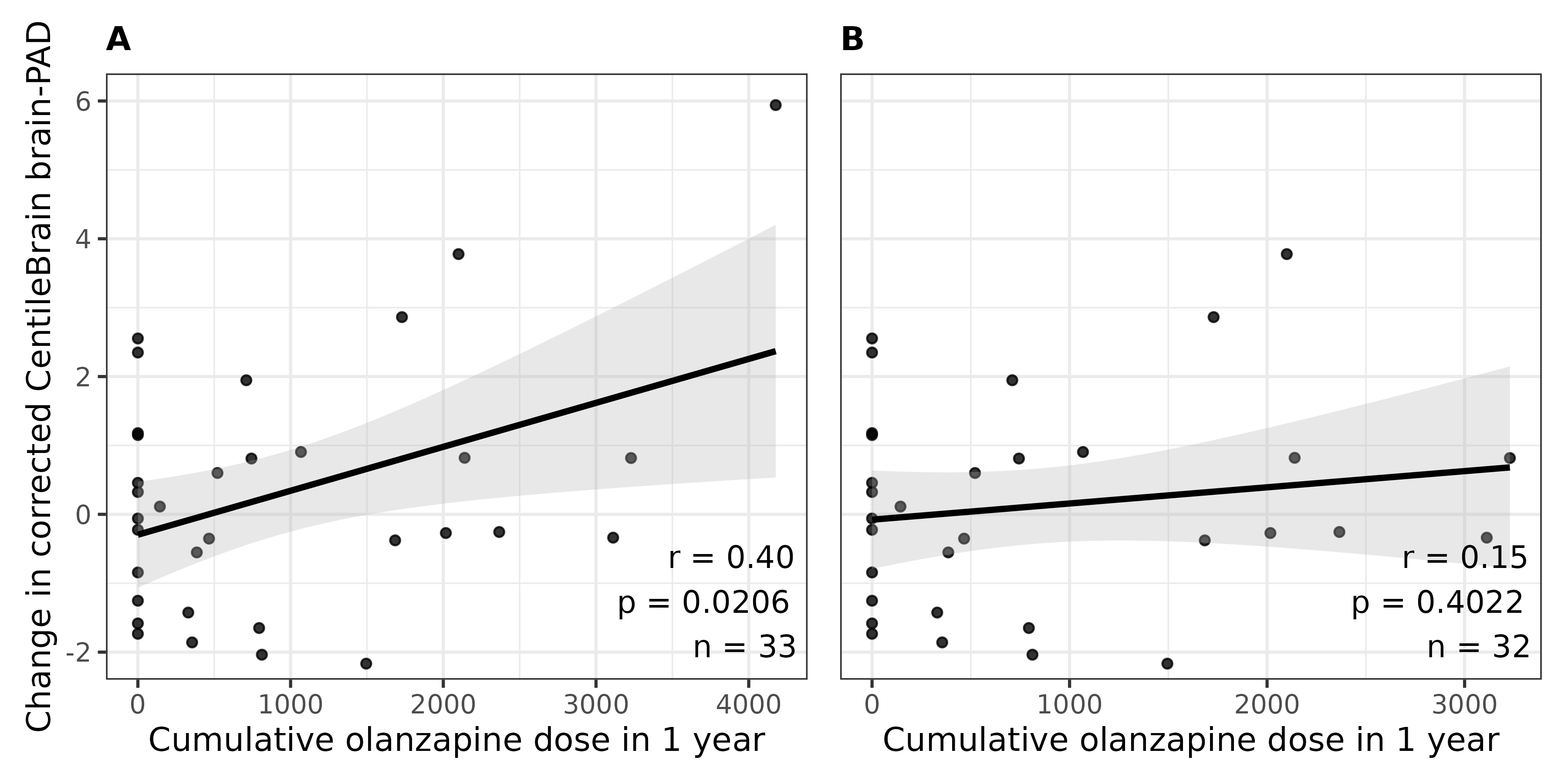

Supplementary figure 5. Longitudinal association between olanzapine exposure and brain aging. In panel A) Scatter plot that includes the outlier. B) Scatter plot without the outlier. Pearson correlation (r), uncorrected p-value (p) and sample size (n) can be seen in the right bottom corner of each panel.
